# Analysis of 200,000 exome-sequenced UK Biobank subjects implicates genes involved in increased and decreased risk of hypertension

**DOI:** 10.1101/2021.02.10.21251503

**Authors:** David Curtis

## Abstract

**Background:** Previous analyses have identified common variants along with some specific genes and rare variants which are associated with risk of hypertension but much remains to be discovered.

**Methods and Results:** Exome-sequenced UK Biobank participants were phenotyped based on having a diagnosis of hypertension or taking anti-hypertensive medication to produce a sample of 66,123 cases and 134,504 controls. Variants with minor allele frequency (MAF) < 0.01 were subjected to a gene-wise weighted burden analysis, with higher weights assigned to variants which are rarer and/or predicted to have more severe effects. Of 20,384 genes analysed, two genes were exome-wide significant, *DNMT3A* and *FES*. Also strongly implicated were *GUCY1A1* and *GUCY1B1*, which code for the subunits of soluble guanylate cyclase. There was further support for the previously reported effects of variants in *NPR1* and protective effects of variants in *DBH*. An inframe deletion in *CACNA1D* with MAF = 0.005, rs72556363, is associated with modestly increased risk of hypertension. Other biologically plausible genes highlighted consist of *CSK, AGTR1, ZYX* and *PREP*. All variants implicated were rare and cumulatively they are not predicted to make a large contribution to the population risk of hypertension.

**Conclusions:** This approach confirms and clarifies previously reported findings and also offers novel insights into biological processes influencing hypertension risk, potentially facilitating the development of improved therapeutic interventions. This research has been conducted using the UK Biobank Resource.

## Introduction

Hypertension is an important risk factor for disease which has a heritable component and a recent large genome wide association study of common variant effects identified 901 loci with enrichment in relevant tissues (blood vessels, heart, adrenal tissue and adipose tissue) and pathways (angiotensinogen, calcium channels, progesterone, natriuretic peptide receptor, angiotensin converting enzyme, angiotensin receptors and endothelin receptors) (Evangelou et al., 2018). Selection pressures tend to mean that common variants individually have small effect sizes and it can be difficult to interpret their biological effects (Wang and Wang, 2018). By contrast, rare variants can potentially have large effect sizes and clear biological mechanisms as exemplified by a number of monogenic causes of hypertension such as congenital adrenal hyperplasia, familial hyperaldosteronism and pseudohypoaldosteronism, which can be caused by variants in *CYP11B1, CYP11B2, WNK1, WNK4, KLHL3, CUL3, SCNN1B, SCNN1G, CYP17A1, HSD11B2, NR3C2* and *KCNJ5* (Ehret and Caulfield, 2013). Additionally, variants in *CACNA1H, CACNA1D* and *CLCN2* have now also been identified as causes of familial hyperaldosteronism while somatic mutations in *ATP1A1* or *ATP2B3* can produce aldosterone-producing adrenal adenomas with consequent hypertension (Scholl et al., 2018). Recessively acting variants in *GUCY1A1* (previously labelled *GUCY1A3*) can cause moyamoya disease and two unrelated subjects with moyamoya disease who also had achalasia and hypertension were found to have compound heterozygote variants in this gene (Wallace et al., 2016). A more recent study has shown that moyamoya disease is itself a risk factor for hypertension (Lee et al., 2020). Using hypertension or blood pressure as phenotypes, gene-based analyses aggregating rare, nonsynonymous variants implicated *PTMT1, DBH* and *NPR1* in a large meta-analysis and also showed that the minor allele of a rare, nonsynonymous variant in *DBH*, rs3025380, was associated with lower blood pressure (Liu et al., 2016). Another study reported that three individual nonsynonymous variants in *NPR1* were associated with increased (rs35479618 and rs116245325) and decreased (rs61757359) blood pressure and showed that this could be explained by the effects of these variants on guanylate cyclase activity (Vandenwijngaert et al., 2019). *KDM1A* codes for LSD1, which removes methyl groups from the methylated lysine 4 residue of histone 3 (H3K4), and there have been reports of association between variants in *KDM1A* and salt-sensitive hypertension in humans, while heterozygous *Lsd1* knock-out mice have salt-sensitive hypertension with increased aldosterone production (Huang et al., 2019; Pojoga et al., 2011; Williams et al., 2012).

The growing availability of sequence data means that it may become possible to study the wider effects of rare, functional variants in the general population. This may implicate novel genes or may demonstrate a wider role for genes already implicated in severe familial disorders. Exome sequence data is now available for 200,000 of the 500,000 UK Biobank subjects (Szustakowski et al., 2020). We have recently analysed this in order to illuminate the effect of rare, coding variants on susceptibility to hyperlipidaemia and a number of other common traits with complex inheritance and we now apply the same approach to study the contribution of rare variants to risk of developing hypertension (Curtis, 2021a).

## Methods

The UK Biobank dataset was downloaded along with the variant call files for 200,632 subjects who had undergone exome-sequencing and genotyping by the UK Biobank Exome Sequencing Consortium using the GRCh38 assembly with coverage 20X at 95.6% of sites on average (Szustakowski et al., 2020). UK Biobank had obtained ethics approval from the North West Multi-centre Research Ethics Committee which covers the UK (approval number: 11/NW/0382) and had obtained informed consent from all participants. The UK Biobank approved an application for use of the data (ID 51119) and ethics approval for the analyses was obtained from the UCL Research Ethics Committee (11527/001). All variants were annotated using the standard software packages VEP, PolyPhen and SIFT (Adzhubei et al., 2013; Kumar et al., 2009; McLaren et al., 2016). To obtain population principal components reflecting ancestry, version 2.0 of *plink* (https://www.cog-genomics.org/plink/2.0/) was run with the options *--maf 0*.*1 --pca 20 approx* (Chang et al., 2015; Galinsky et al., 2016).

The UK Biobank sample contains 503,317 subjects of whom 94.6% are of white ethnicity. As we have discussed previously, it has become standard practice for investigators to simply discard data from participants with other ancestries and we regard this as regrettable (Curtis, 2021b). We demonstrated that if population principal components are included as covariates then it is possible to include all participants, regardless of ancestry, in the type of weighted burden analysis described here without inflation of the test statistic.

To define cases, a similar approach was used as was previously implemented for the investigation of hyperlipidaemia and T2D (Curtis, 2021a, 2021c, 2019). The hypertension phenotype was determined from four sources in the dataset: self-reported diagnosis recorded as hypertension or essential hypertension; reporting taking medication for high blood pressure; reporting taking any of a list of named medications commonly used to treat high blood pressure (https://www.nhs.uk/conditions/high-blood-pressure-hypertension/); having an ICD10 diagnosis of essential hypertension, hypertensive heart disease or hypertensive renal disease in hospital records or as a cause of death. Subjects in any of these categories were deemed to be cases with hypertension while all other subjects were taken to be controls.

The same analytic methods as had been used previously were applied, with the description repeated here for the reader’s convenience. The SCOREASSOC program was used to carry out a weighted burden analysis to test whether, in each gene, sequence variants which were rarer and/or predicted to have more severe functional effects occurred more commonly in cases than controls. Attention was restricted to rare variants with minor allele frequency (MAF) <= 0.01 in both cases and controls. As previously described, variants were weighted by overall MAF so that variants with MAF=0.01 were given a weight of 1 while very rare variants with MAF close to zero were given a weight of 10 (Curtis, 2021b). Variants were also weighted according to their functional annotation using the GENEVARASSOC program, which was used to generate input files for weighted burden analysis by SCOREASSOC (Curtis, 2016, 2012). The weights were informed from the analysis of the effects of different categories of variant in *LDLR* on hyperlipidaemia risk (Curtis, 2021a). Variants predicted to cause complete loss of function (LOF) of the gene were assigned a weight of 100. Nonsynonymous variants were assigned a weight of 5 but if PolyPhen annotated them as possibly or probably damaging then 5 or 10 was added to this and if SIFT annotated them as deleterious then 20 was added. In order to allow exploration of the effects of different types of variant on disease risk the variants were also grouped into broader categories to be used in multivariate analyses as described below. The full set of weights and categories is displayed in Table 1. As described previously, the weight due to MAF and the weight due to functional annotation were multiplied together to provide an overall weight for each variant. Variants were excluded if there were more than 10% of genotypes missing in the controls or if the heterozygote count was smaller than both homozygote counts in the controls. If a subject was not genotyped for a variant then they were assigned the subject-wise average score for that variant. For each subject a gene-wise weighted burden score was derived as the sum of the variant-wise weights, each multiplied by the number of alleles of the variant which the given subject possessed. For variants on the X chromosome, hemizygous males were treated as homozygotes.

**Table 1.** The table shows the weight which was assigned to each type of variant as annotated by VEP, Polyphen and SIFT as well as the broad categories which were used for multivariate analyses of variant effects (Adzhubei et al., 2013; Kumar et al., 2009; McLaren et al., 2016).

| **VEP / SIFT / Polyphen annotation** | **Weight** | **Category** |
| --- | --- | --- |
| intergenic_variant | 0 | Unused |
| feature_truncation | 0 | Intronic, etc. |
| regulatory_region_variant | 0 | Intronic, etc. |
| feature_elongation | 0 | Intronic, etc. |
| regulatory_region_amplification | 1 | Intronic, etc. |
| regulatory_region_ablation | 1 | Intronic, etc. |
| TF_binding_site_variant | 1 | Intronic, etc. |
| TFBS_amplification | 1 | Intronic, etc. |
| TFBS_ablation | 1 | Intronic, etc. |
| downstream_gene_variant | 0 | Intronic, etc. |
| upstream_gene_variant | 0 | Intronic, etc. |
| non_coding_transcript_variant | 0 | Intronic, etc. |
| NMD_transcript_variant | 0 | Intronic, etc. |
| intron_variant | 0 | Intronic, etc. |
| non_coding_transcript_exon_variant | 0 | Intronic, etc. |
| 3_prime_UTR_variant | 1 | 3 prime UTR |
| 5_prime_UTR_variant | 1 | 5 prime UTR |
| mature_miRNA_variant | 5 | Unused |
| coding_sequence_variant | 0 | Unused |
| synonymous_variant | 0 | Synonymous |
| stop_retained_variant | 5 | Unused |
| incomplete_terminal_codon_variant | 5 | Unused |
| splice_region_variant | 1 | Splice region |
| protein_altering_variant | 5 | Protein altering |
| missense_variant | 5 | Protein altering |
| inframe_deletion | 10 | InDel, etc |
| inframe_insertion | 10 | InDel, etc |
| transcript_amplification | 10 | InDel, etc |
| start_lost | 10 | Unused |
| stop_lost | 10 | Unused |
| frameshift_variant | 100 | Disruptive |
| stop_gained | 100 | Disruptive |
| splice_donor_variant | 100 | Splice site variant |
| splice_acceptor_variant | 100 | Splice site variant |
| transcript_ablation | 100 | Disruptive |
| SIFT deleterious | 20 | Deleterious |
| PolyPhen possibly damaging | 5 | Possibly damaging |
| PolyPhen probably damaging | 10 | Probably damaging |

For each gene, logistic regression analysis was carried out including the first 20 population principal components and sex as covariates and a likelihood ratio test was performed comparing the likelihoods of the models with and without the gene-wise burden score. The statistical significance was summarised as a signed log p value (SLP), which is the log base 10 of the p value given a positive sign if the score is higher in cases and negative if it is higher in controls.

Gene set analyses were carried out as before using the 1454 “all GO gene sets, gene symbols” pathways as listed in the file *c5*.*all*.*v5*.*0*.*symbols*.*gmt* downloaded from the Molecular Signatures Database at http://www.broadinstitute.org/gsea/msigdb/collections.jsp (Subramanian et al., 2005). For each set of genes, the natural logs of the gene-wise p values were summed according to Fisher’s method to produce a chi-squared statistic with degrees of freedom equal to twice the number of genes in the set. The p value associated with this chi-squared statistic was expressed as a minus log10 p (MLP) as a test of association of the set with the hyperlipidaemia phenotype.

For selected genes, additional analyses were carried out to clarify the contribution of different categories of variant. As described previously, logistic regression analyses were performed on the counts of the separate categories of variant as listed in Table 1, again including principal components and sex as covariates, to estimate the effect size for each category (Curtis, 2021a). The odds ratios (ORs) associated with each category were estimated along with their standard errors and the Wald statistic was used to obtain a p value, except for categories in which variants occurred fewer than 50 times in which case Fisher’s exact test was applied to the variant counts. The associated p value was converted to an SLP, again with the sign being positive if the OR was greater than 1, indicating that variants in that category tended to increase risk.

Data manipulation and statistical analyses were performed using GENEVARASSOC, SCOREASSOC and R (R Core Team, 2014).

## Results

There were 66,123 cases and 134,504 controls. There were 20,384 genes for which there were qualifying variants. Given that there were 20,384 informative genes, the critical threshold for the absolute value of the SLP to declare a result as formally statistically significant is -log10(0.05/20384) = 5.61 and this was achieved by two genes, *DNMT3A* (SLP = 8.21) and *FES* (SLP = 6.10). The quantile-quantile (QQ) plot for the SLPs obtained for all genes except *DNMT3A* is shown in Figure 1. This shows that the test appears to be well-behaved and conforms well with the expected distribution. Omitting the genes with the 100 highest and 100 lowest SLPs, which might be capturing a real biological effect, the gradient for positive SLPs is 1.096 with intercept at 0.006 and the gradient for negative SLPs is 1.080 with intercept at 0.02, indicating only modest inflation of the test statistic in spite of the fact that participants of all ancestries are included.

**Figure 1.**
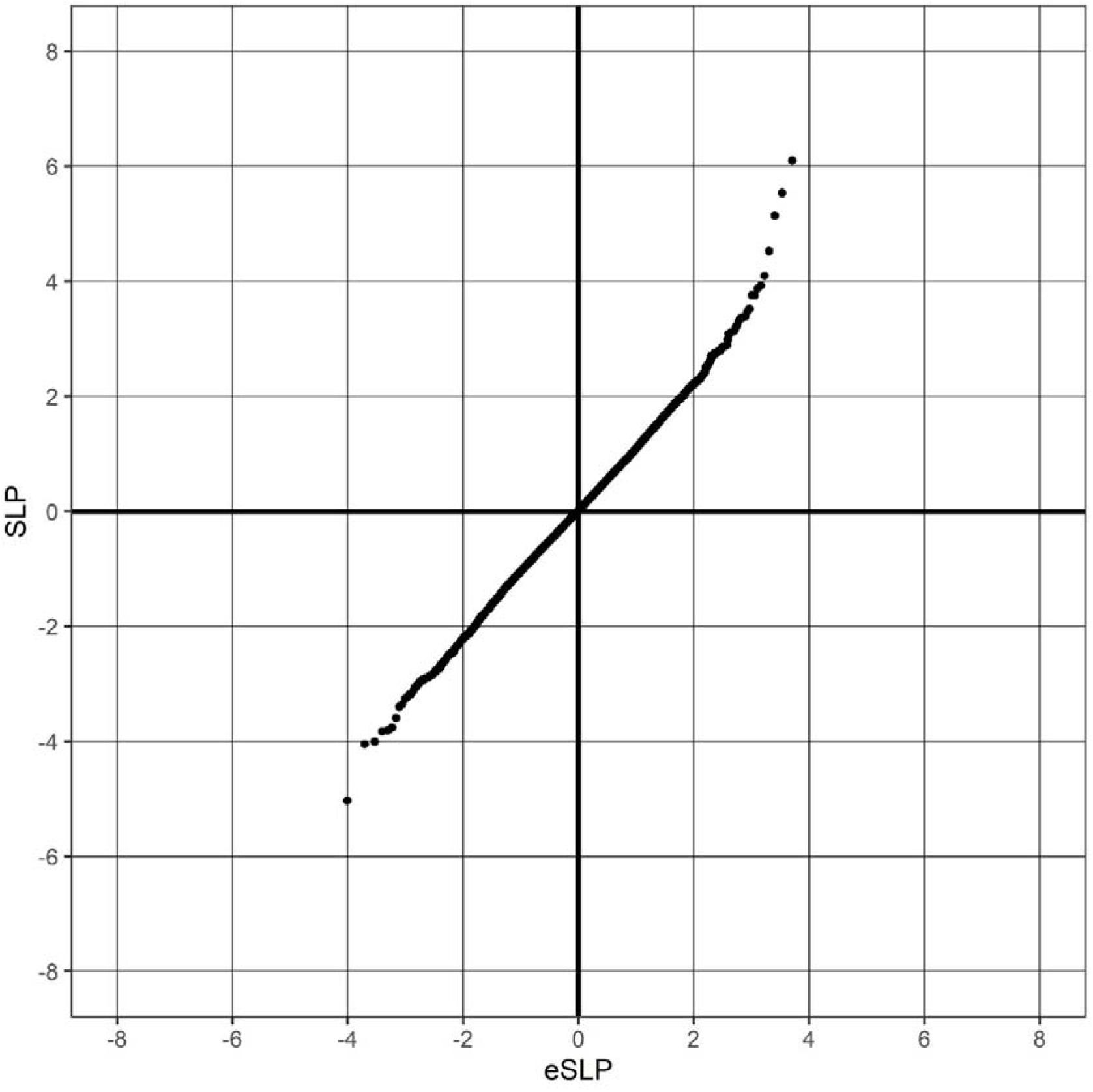
QQ plot of SLPs obtained for weighted burden analysis of association with hypertension showing observed against expected SLP for each gene, omitting results for *DNMT3A*, which has SLP = 8.21.

Table 2 shows all the genes achieving SLP with absolute value greater than 3, equivalent to an uncorrected p value of 0.001. Given that 20,384 genes were tested, one would expect that by chance about 20 would reach this level of significance whereas in fact there are 42. Thus it is possible that some of these highly ranked genes do demonstrate a biological signal which fails to reach statistical significance after correction for multiple testing. For *NPR1*, the analysis was repeated excluding data from the previously reported individually significant variants rs35479618, rs116245325 and rs61757359. This resulted in a reduction of the SLP from 5.14 to 4.38. For *DBH*, the analysis was repeated without rs3025380 and this resulted in a change in SLP from −3.40 to −2.11. The full list of results for all genes is provided in Supplementary Table S1.

**Table 2.** Genes with absolute value of SLP exceeding 3 or more (equivalent to p<0.001) for test of association of weighted burden score with hypertension.

| **Symbol** | **SLP** | **Name** |
| --- | --- | --- |
| *DNMT3A* | 8.21 | DNA Methyltransferase 3 Alpha |
| *FES* | 6.10 | FES Proto-Oncogene, Tyrosine Kinase |
| *GUCY1A1* | 5.54 | Guanylate Cyclase 1 Soluble Subunit Alpha 1 |
| *NPR1* | 5.14 | Natriuretic Peptide Receptor 1 |
| *MAG* | 4.52 | Myelin Associated Glycoprotein |
| *SMAD6* | 4.10 | SMAD Family Member 6 |
| *GUCY1B1* | 3.93 | Guanylate Cyclase 1 Soluble Subunit Beta 1 |
| *SKIV2L* | 3.87 | Ski2 Like RNA Helicase |
| *ASXL1* | 3.75 | ASXL Transcriptional Regulator 1 |
| *MYO10* | 3.75 | Myosin X |
| *PTGER3* | 3.52 | Prostaglandin E Receptor 3 |
| *POLR3H* | 3.47 | RNA Polymerase III Subunit H |
| *IFT172* | 3.39 | Intraflagellar Transport 172 |
| *LITAF* | 3.37 | Lipopolysaccharide Induced TNF Factor |
| *CSK* | 3.37 | C-Terminal Src Kinase |
| *SNX27* | 3.34 | Sorting Nexin 27 |
| *TEK* | 3.31 | EK Receptor Tyrosine Kinase |
| *PHF21A* | 3.23 | PHD Finger Protein 21A |
| *LOC102723675* | 3.21 | Uncharacterized LOC102723675 |
| *DXO* | 3.14 | Decapping Exoribonuclease |
| *RHBDL3* | 3.12 | Rhomboid Like 3 |
| *ASB7* | 3.12 | Ankyrin Repeat And SOCS Box Containing 7 |
| *MPP2* | 3.11 | Membrane Palmitoylated Protein 2 |
| *ZNF549* | 3.10 | Zinc Finger Protein 549 |
| *MAP3K14-AS1* | 3.08 | MAP3K14 Antisense RNA 1 |
| *ZNF600* | -3.01 | Zinc Finger Protein 600 |
| *SIRT2* | -3.05 | Sirtuin 2 |
| *PARGP1* | -3.05 | Poly(ADP-Ribose) Glycohydrolase Pseudogene 1 |
| *ZNF226* | -3.13 | Zinc Finger Protein 226 |
| *LOC105375153* | -3.18 | Uncharacterized LOC105375153 |
| *INPPL1* | -3.19 | Inositol Polyphosphate Phosphatase Like 1 |
| *FNDC8* | -3.23 | Fibronectin Type III Domain Containing 8 |
| *KRT82* | -3.26 | Keratin 82 |
| *GOLGA4* | -3.36 | Golgin A4 |
| *DBH* | -3.40 | Dopamine Beta-Hydroxylase |
| *ELOF1* | -3.59 | Elongation Factor 1 Homolog |
| *AGTR1* | -3.77 | Angiotensin II Receptor Type 1 |
| *SIPA1* | -3.81 | Signal-Induced Proliferation-Associated 1 |
| *ZYX* | -3.83 | Zyxin |
| *ZFAT* | -4.00 | Zinc Finger And AT-Hook Domain Containing |
| *OSTF1* | -4.05 | Osteoclast Stimulating Factor 1 |
| *PREP* | -5.03 | Prolyl Endopeptidase |

In order to see if any additional genes were highlighted by analysing gene sets, gene set analysis was performed as described above after first dividing the gene-wise log p values by the average inflation factor of 1.09 before combining them using Fisher’s method. Given that 1,454 sets were tested, a critical MLP to achieve to declare results significant after correction for multiple testing would be log10(1454*20) = 4.46 and this was achieved by just one set, labelled CHROMATIN. This set contains 35 genes, including *DNMT3A*. The full results of the gene set analyses are listed in Supplementary Table S2. Four other sets achieved SLP > 3 and these and for these all genes with an absolute value SLP > 1.3 (equivalent to p < 0.05) were listed. All of these sets also contained *DNMT3A* and none of the other nominally significant genes appeared to be obviously relevant to hypertension. These results are presented in Supplementary Table S3.

For the genes listed in Table 2 which appeared to be of interest, additional multivariate analyses were performed to elucidate the contribution to the overall result from different categories of variant. The results of this analysis for *DNMT3A* are shown in Table 3A. From this it can be seen that the signal comes from disruptive and splice site variants which are predicted to cause LOF and which are between them associated with an OR of about 1.9. However, variants annotated as probably damaging by PolyPhen are also commoner in cases and are associated with an OR of 1.5. Table 3B shows that the result for *FES* is driven by a small number of disruptive variants which are commoner in cases with OR of 2.8. It is striking that both *GUCY1A1* and *GUCY1B1* are ranked among the top 7 genes since they code for subunits of the same guanylate cyclase and their results are shown in full in Table 3C and 3D. This shows that while the effect for *GUCY1A1* is driven by LOF variants these are very rare in *GUCY1B1* and for this gene there seems to be an additional contribution from an excess of 5 prime UTR variants among cases. These occur at 44 different locations and detailed inspection of the output file showed these all to be individually rare, such that there was no single variant which could be seen to making a significant contribution to the overall effect. For the other genes thought to be of interest, Table 4 provides a summary of the results for LOF variants along with any other variant category individually significant at p < 0.05. Full results for analyses of variant categories are presented in Supplementary Table S4.

**Table 3.** Results from logistic regression analysis showing the effects on risk of hypertension of different categories of variant within *DNMT3A, FES, GUCY1A1* and *GUCY1B1*. Odds ratios for each category are estimated including principal components and sex as covariates. The SLP is also obtained from this multivariate analysis except when there are fewer than 50 variants in a category, when Fisher’s exact test is used instead. **Table 3A** Results for *DNMT3A*.

| Category | Number of separate variants | Total count in controls | Mean count in controls | Total count in cases | Mean count in cases | OR (95% confidence interval) | SLP |
| --- | --- | --- | --- | --- | --- | --- | --- |
| Intronic, etc | 1417 | 15625 | 0.116168 | 7859 | 0.118854 | 0.98 (0.95 - 1.00) | 1.04 |
| 5 prime UTR | 71 | 395 | 0.002937 | 187 | 0.002828 | 0.96 (0.80 - 1.15) | -0.20 |
| Synonymous | 157 | 2427 | 0.018044 | 1342 | 0.020296 | 1.04 (0.97 - 1.11) | 0.55 |
| Splice region | 63 | 339 | 0.002520 | 206 | 0.003115 | 1.10 (0.92 - 1.32) | 0.54 |
| 3 prime UTR | 19 | 37 | 0.000275 | 10 | 0.000151 | 0.57 (0.28 - 1.16) | -0.92 |
| Protein altering | 227 | 1310 | 0.009739 | 729 | 0.011025 | 1.08 (0.88 - 1.33) | 0.36 |
| InDel, etc | 7 | 13 | 0.000097 | 10 | 0.000151 | 1.62 (0.69 - 3.77) | 0.56 |
| Disruptive | 53 | 76 | 0.000565 | 67 | 0.001013 | 1.86 (1.33 - 2.60) | 3.62 |
| Splice site variant | 21 | 43 | 0.000320 | 39 | 0.000590 | 1.87 (1.20 - 2.92) | 2.31 |
| Deleterious | 119 | 1033 | 0.007680 | 577 | 0.008726 | 0.98 (0.75 - 1.28) | 0.06 |
| Possibly damaging | 39 | 659 | 0.004899 | 339 | 0.005127 | 0.99 (0.78 - 1.26) | 0.04 |
| Probably damaging | 68 | 185 | 0.001375 | 143 | 0.002163 | 1.50 (1.11 - 2.03) | 2.18 |

**Table 3B.**
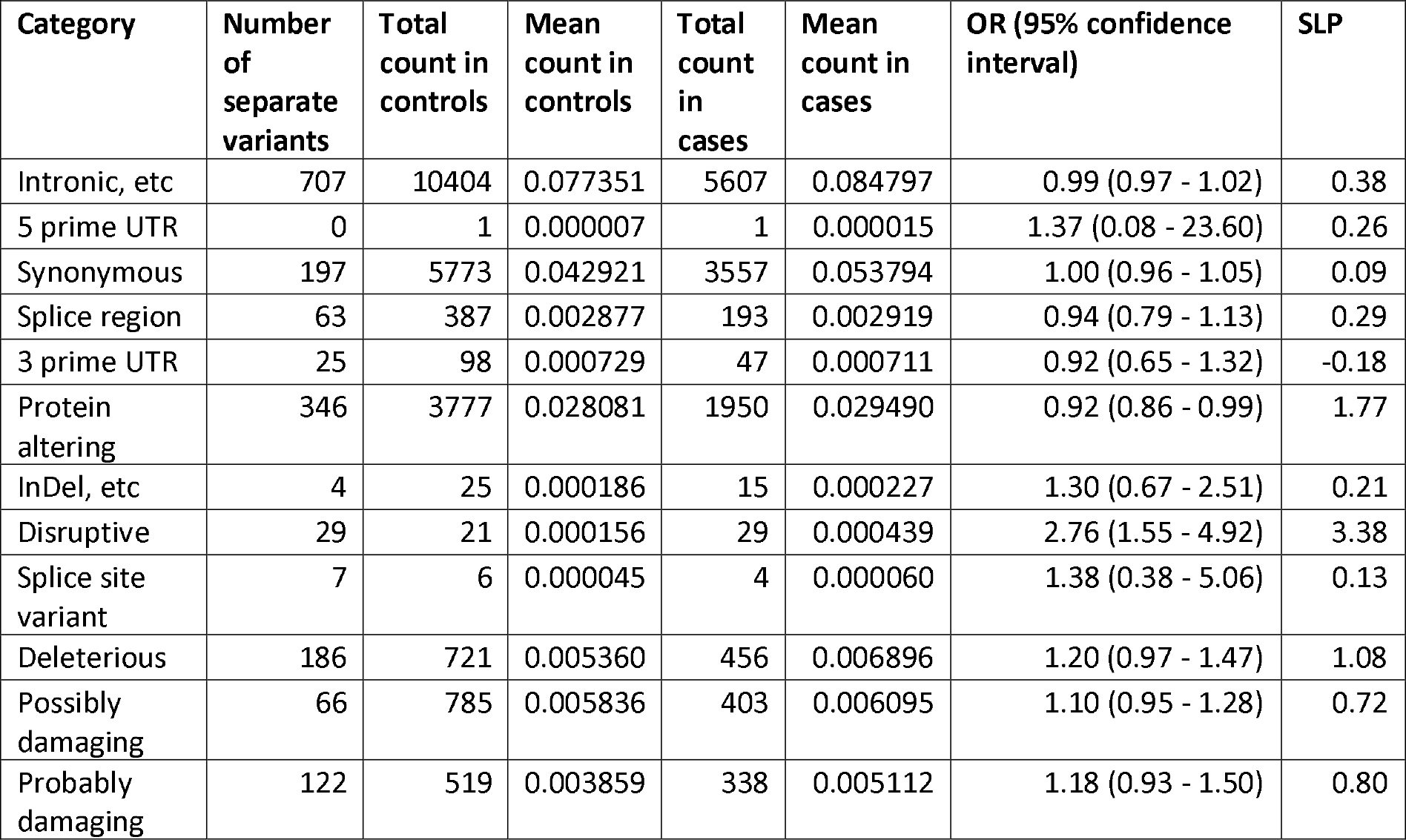
Results for *FES*.

| Category | Number of separate variants | Total count in controls | Mean count in controls | Total count in cases | Mean count in cases | OR (95% confidence interval) | SLP |
| --- | --- | --- | --- | --- | --- | --- | --- |
| Intronic, etc | 707 | 10404 | 0.077351 | 5607 | 0.084797 | 0.99 (0.97 - 1.02) | 0.38 |
| 5 prime UTR | 0 | 1 | 0.000007 | 1 | 0.000015 | 1.37 (0.08 - 23.60) | 0.26 |
| Synonymous | 197 | 5773 | 0.042921 | 3557 | 0.053794 | 1.00 (0.96 - 1.05) | 0.09 |
| Splice region | 63 | 387 | 0.002877 | 193 | 0.002919 | 0.94 (0.79 - 1.13) | 0.29 |
| 3 prime UTR | 25 | 98 | 0.000729 | 47 | 0.000711 | 0.92 (0.65 - 1.32) | -0.18 |
| Protein altering | 346 | 3777 | 0.028081 | 1950 | 0.029490 | 0.92 (0.86 - 0.99) | 1.77 |
| InDel, etc | 4 | 25 | 0.000186 | 15 | 0.000227 | 1.30 (0.67 - 2.51) | 0.21 |
| Disruptive | 29 | 21 | 0.000156 | 29 | 0.000439 | 2.76 (1.55 - 4.92) | 3.38 |
| Splice site variant | 7 | 6 | 0.000045 | 4 | 0.000060 | 1.38 (0.38 - 5.06) | 0.13 |
| Deleterious | 186 | 721 | 0.005360 | 456 | 0.006896 | 1.20 (0.97 - 1.47) | 1.08 |
| Possibly damaging | 66 | 785 | 0.005836 | 403 | 0.006095 | 1.10 (0.95 - 1.28) | 0.72 |
| Probably damaging | 122 | 519 | 0.003859 | 338 | 0.005112 | 1.18 (0.93 - 1.50) | 0.80 |

**Table 3C.** Results for *GUCY1A1*.

| Category | Number of separate variants | Total count in controls | Mean count in controls | Total count in cases | Mean count in cases | OR (95% confidence interval) | SLP |
| --- | --- | --- | --- | --- | --- | --- | --- |
| Intronic, etc | 355 | 9682 | 0.071983 | 5306 | 0.080244 | 1.02 (0.98 - 1.06) | 0.57 |
| 5 prime UTR | 27 | 1610 | 0.011970 | 864 | 0.013067 | 1.06 (0.97 - 1.15) | 0.77 |
| Synonymous | 114 | 4002 | 0.029754 | 2438 | 0.036871 | 1.02 (0.96 - 1.08) | 0.24 |
| Splice region | 18 | 35 | 0.000260 | 22 | 0.000333 | 1.25 (0.72 - 2.15) | 0.37 |
| 3 prime UTR | 16 | 64 | 0.000476 | 39 | 0.000590 | 1.30 (0.86 - 1.96) | 0.70 |
| Protein altering | 281 | 2538 | 0.018869 | 1635 | 0.024727 | 1.00 (0.92 - 1.09) | 0.00 |
| InDel, etc | 2 | 2 | 0.000015 | 0 | 0.000000 |  | 0.00 |
| Disruptive | 29 | 35 | 0.000260 | 39 | 0.000590 | 2.41 (1.51 - 3.85) | 3.76 |
| Splice site variant | 5 | 13 | 0.000097 | 12 | 0.000181 | 1.84 (0.82 - 4.11) | 0.87 |
| Deleterious | 122 | 395 | 0.002937 | 242 | 0.003660 | 1.11 (0.81 - 1.52) | 0.31 |
| Possibly damaging | 41 | 294 | 0.002186 | 158 | 0.002389 | 1.04 (0.83 - 1.31) | 0.14 |
| Probably damaging | 92 | 285 | 0.002119 | 175 | 0.002647 | 1.12 (0.79 - 1.61) | 0.29 |

**Table 3D.** Results for *GUCY1B1*.

| Category | Number of separate variants | Total count in controls | Mean count in controls | Total count in cases | Mean count in cases | OR (95% confidence interval) | SLP |
| --- | --- | --- | --- | --- | --- | --- | --- |
| Intronic, etc | 599 | 5482 | 0.040757 | 3271 | 0.049468 | 1.01 (0.97 - 1.06) | 0.22 |
| 5 prime UTR | 44 | 163 | 0.001212 | 114 | 0.001724 | 1.37 (1.07 - 1.75) | 1.94 |
| Synonymous | 96 | 3833 | 0.028497 | 1862 | 0.028160 | 0.99 (0.93 - 1.05) | -0.17 |
| Splice region | 38 | 107 | 0.000796 | 53 | 0.000802 | 1.00 (0.71 - 1.40) | 0.01 |
| 3 prime UTR | 20 | 119 | 0.000885 | 44 | 0.000665 | 0.77 (0.54 - 1.10) | -0.86 |
| Protein altering | 187 | 990 | 0.007360 | 545 | 0.008242 | 1.06 (0.94 - 1.20) | 0.46 |
| InDel, etc | 1 | 4 | 0.000030 | 0 | 0.000000 |  | -0.51 |
| Disruptive | 12 | 10 | 0.000074 | 11 | 0.000166 | 2.06 (0.86 - 4.97) | 1.18 |
| Splice site variant | 10 | 7 | 0.000052 | 12 | 0.000181 | 3.43 (1.32 - 8.94) | 1.92 |
| Deleterious | 87 | 202 | 0.001502 | 128 | 0.001936 | 1.09 (0.75 - 1.56) | 0.19 |
| Possibly damaging | 29 | 68 | 0.000506 | 44 | 0.000665 | 0.98 (0.65 - 1.48) | 0.03 |
| Probably damaging | 59 | 109 | 0.000810 | 75 | 0.001134 | 1.21 (0.76 - 1.91) | 0.38 |

**Table 4.** Summary results of variant category analysis for additional genes of interest. Results are shown for disruptive and splice site variants and for any other variant categories significant at p < 0.05.

| Gene / Category | Number of separate variants | Total count in controls | Mean count in controls | Total count in cases | Mean count in cases | OR (95% confidence interval) | SLP |
| --- | --- | --- | --- | --- | --- | --- | --- |
| *NPR1* |  |  |  |  |  |  |  |
| Disruptive | 24 | 50 | 0.000372 | 44 | 0.000665 | 1.75 (1.15 - 2.65) | 2.14 |
| Splice site variant | 7 | 10 | 0.000074 | 5 | 0.000076 | 1.07 (0.35 - 3.22) | 0.00 |
| Probably damaging | 105 | 281 | 0.002089 | 195 | 0.002949 | 1.32 (1.05 - 1.66) | 1.81 |
| *SMAD6* |  |  |  |  |  |  |  |
| Synonymous | 193 | 3378 | 0.025114 | 1558 | 0.023562 | 0.94 (0.88 - 1.00) | -1.31 |
| Disruptive | 103 | 92 | 0.000684 | 68 | 0.001028 | 1.48 (1.07 - 2.03) | 1.84 |
| Splice site variant | 7 | 4 | 0.000030 | 3 | 0.000045 | 1.65 (0.36 - 7.68) | 0.16 |
| Possibly damaging | 74 | 208 | 0.001546 | 137 | 0.002072 | 1.29 (1.02 - 1.63) | 1.55 |
| *IFT172* |  |  |  |  |  |  |  |
| Protein altering | 711 | 7499 | 0.055753 | 4090 | 0.061854 | 1.07 (1.01 - 1.13) | 1.82 |
| Disruptive | 58 | 94 | 0.000699 | 65 | 0.000983 | 1.39 (1.01 - 1.91) | 1.40 |
| Splice site variant | 22 | 72 | 0.000535 | 34 | 0.000514 | 0.98 (0.64 - 1.49) | -0.04 |
| *CSK* |  |  |  |  |  |  |  |
| Splice region | 38 | 969 | 0.007204 | 782 | 0.011826 | 1.20 (1.01 - 1.42) | 1.47 |
| Disruptive | 3 | 2 | 0.000015 | 1 | 0.000015 | 0.93 (0.08 - 10.86) | 0.00 |
| Splice site variant | 1 | 1 | 0.000007 | 1 | 0.000015 | 2.02 (0.12 - 34.90) | 0.26 |
| Deleterious | 36 | 193 | 0.001435 | 182 | 0.002752 | 1.45 (1.07 - 1.97) | 1.83 |
| *DBH* |  |  |  |  |  |  |  |
| Disruptive | 27 | 71 | 0.000528 | 28 | 0.000423 | 0.83 (0.53 - 1.29) | -0.40 |
| Splice site variant | 8 | 544 | 0.004044 | 256 | 0.003872 | 0.96 (0.82 - 1.12) | -0.24 |
| Deleterious | 186 | 5384 | 0.040029 | 2343 | 0.035434 | 0.86 (0.76 - 0.98) | -1.65 |
| *AGTR1* |  |  |  |  |  |  |  |
| Disruptive | 25 | 64 | 0.000476 | 13 | 0.000197 | 0.41 (0.22 - 0.75) | -2.46 |
| Splice site variant | 0 | 0 | 0.000000 | 0 | 0.000000 |  | 0.00 |
| *ZYX* |  |  |  |  |  |  |  |
| Synonymous | 149 | 3694 | 0.027464 | 2162 | 0.032697 | 1.05 (1.00 - 1.11) | 1.32 |
| Disruptive | 27 | 58 | 0.000431 | 9 | 0.000136 | 0.32 (0.16 - 0.67) | -2.76 |
| Splice site variant | 6 | 10 | 0.000074 | 4 | 0.000060 | 0.88 (0.27 - 2.89) | 0.00 |
| *PREP* |  |  |  |  |  |  |  |
| Disruptive | 18 | 20 | 0.000149 | 5 | 0.000076 | 0.50 (0.18 - 1.37) | -0.69 |
| Splice site variant | 3 | 3 | 0.000022 | 0 | 0.000000 |  | -0.26 |

Of the previously implicated genes listed in the Introduction, aside from *GUCY1A1* only the following were significant at p < 0.05: *CYP11B1* (SLP = 1.63), *NR3C2* (SLP = 1.67) and *CACNA1H* (SLP = −2.05). Analyses of the variant categories were carried out for these genes and a summary of the results is shown in Table 5, which provides the results for LOF variants and for other categories which were significant at p < 0.05. These are mostly unremarkable and it is difficult to draw firm conclusions although a few results are worth noting. For most genes LOF variants are very rare so that one cannot gain a clear estimate of their effect. However the results for *WNK1, WNK4, CLCN2* and *ATP2B3* suggest that LOF variants in these genes do not have a very major effect on risk of hypertension. The most striking result is that for *CACNA1D* the InDel category produces SLP = 8.10 with OR = 1.30. InDel variants occur at 9 locations in this gene and inspection of the detailed results revealed that 8 of these are very rare so the result reflects the effect of a single inframe deletion, 3:53808664-CCTT>C. This is rs72556363, which results in the loss of a phenylalanine residue, p.Phe1923del, and according to gnomAD it has allele frequency of 0.0043 in non-Finnish Europeans and is extremely rare or absent from other populations. In the UK Biobank subjects we observe a frequency of 0.0049 in controls and 0.0064 in cases. Full results for analyses of variant categories in these genes are presented in Supplementary Table S5.

**Table 5.** Summary results of variant category analysis for previously implicated genes. Results are shown for disruptive and splice site variants and for any other variant categories significant at p < 0.05.

| Gene / Category | Number of separate variants | Total count in controls | Mean count in controls | Total count in cases | Mean count in cases | OR (95% confidence interval) | SLP |
| --- | --- | --- | --- | --- | --- | --- | --- |
| *CYP11B1* |  |  |  |  |  |  |  |
| Disruptive | 22 | 68 | 0.000506 | 38 | 0.000575 | 1.14 (0.76 - 1.72) | 0.29 |
| Splice site variant | 6 | 21 | 0.000156 | 7 | 0.000106 | 0.71 (0.30 - 1.71) | -0.37 |
| Probably damaging | 93 | 531 | 0.003948 | 340 | 0.005142 | 1.27 (1.03 - 1.57) | 1.60 |
| *CYP11B2* |  |  |  |  |  |  |  |
| Synonymous | 150 | 3649 | 0.027129 | 1830 | 0.027676 | 0.94 (0.88 - 0.99) | -1.72 |
| Protein altering | 268 | 2193 | 0.016304 | 1251 | 0.018919 | 1.13 (1.03 - 1.25) | 2.07 |
| Disruptive | 19 | 37 | 0.000275 | 23 | 0.000348 | 1.28 (0.75 - 2.18) | 0.45 |
| Splice site variant | 7 | 46 | 0.000342 | 21 | 0.000318 | 0.93 (0.55 - 1.59) | -0.10 |
| SIFT | 129 | 957 | 0.007115 | 456 | 0.006896 | 0.74 (0.61 - 0.91) | -2.53 |
| *WNK1* |  |  |  |  |  |  |  |
| Disruptive | 51 | 157 | 0.001167 | 84 | 0.001270 | 1.08 (0.82 - 1.42) | 0.25 |
| Splice site variant | 8 | 31 | 0.000230 | 20 | 0.000302 | 1.16 (0.65 - 2.07) | 0.22 |
| *WNK4* |  |  |  |  |  |  |  |
| Intronic, etc | 576 | 12657 | 0.094101 | 7753 | 0.117251 | 1.04 (1.00 - 1.07) | 1.51 |
| Disruptive | 54 | 301 | 0.002238 | 165 | 0.002495 | 1.12 (0.92 - 1.36) | 0.58 |
| Splice site variant | 18 | 24 | 0.000178 | 8 | 0.000121 | 0.67 (0.29 - 1.51) | -0.34 |
| *KLHL3* |  |  |  |  |  |  |  |
| Disruptive | 5 | 6 | 0.000045 | 1 | 0.000015 | 0.33 (0.04 - 2.85) | -0.36 |
| Splice site variant | 4 | 5 | 0.000037 | 2 | 0.000030 | 0.95 (0.18 - 5.09) | 0.00 |
| *CUL3* |  |  |  |  |  |  |  |
| 5 prime UTR | 41 | 108 | 0.000803 | 85 | 0.001285 | 1.54 (1.15 - 2.06) | 2.51 |
| InDel, etc | 3 | 0 | 0.000000 | 3 | 0.000045 |  | 1.45 |
| Disruptive | 4 | 4 | 0.000030 | 1 | 0.000015 | 0.53 (0.06 - 4.97) | 0.00 |
| *SCNN1B* |  |  |  |  |  |  |  |
| Disruptive | 16 | 31 | 0.000230 | 11 | 0.000166 | 0.73 (0.36 - 1.48) | -0.38 |
| Splice site variant | 8 | 10 | 0.000074 | 7 | 0.000106 | 1.64 (0.61 - 4.42) | 0.35 |
| *SCNN1G* |  |  |  |  |  |  |  |
| InDel, etc | 3 | 68 | 0.000506 | 18 | 0.000272 | 0.54 (0.31 - 0.91) | -1.72 |
| Disruptive | 12 | 14 | 0.000104 | 8 | 0.000121 | 1.16 (0.48 - 2.83) | 0.09 |
| Splice site variant | 4 | 17 | 0.000126 | 9 | 0.000136 | 1.06 (0.46 - 2.43) | 0.08 |
| *CYP17A1* |  |  |  |  |  |  |  |
| Intronic, etc | 293 | 3572 | 0.026557 | 2081 | 0.031472 | 1.06 (1.00 - 1.11) | 1.46 |
| Disruptive | 16 | 57 | 0.000424 | 28 | 0.000423 | 0.96 (0.60 - 1.52) | -0.07 |
| Splice site variant | 2 | 1 | 0.000007 | 2 | 0.000030 | 4.36 (0.37 - 50.99) | 0.59 |
| *HSD11B2* |  |  |  |  |  |  |  |
| Synonymous | 85 | 2500 | 0.018587 | 1581 | 0.023910 | 0.93 (0.86 - 1.00) | -1.37 |
| Splice region | 17 | 803 | 0.005970 | 377 | 0.005701 | 0.86 (0.76 - 0.98) | -1.67 |
| Disruptive | 8 | 16 | 0.000119 | 5 | 0.000076 | 0.66 (0.24 - 1.85) | -0.31 |
| Splice site variant | 4 | 7 | 0.000052 | 4 | 0.000060 | 1.08 (0.30 - 3.80) | 0.12 |
| Deleterious | 91 | 226 | 0.001680 | 118 | 0.001785 | 0.69 (0.50 - 0.96) | -1.61 |
| Probably damaging | 46 | 68 | 0.000506 | 59 | 0.000892 | 2.50 (1.55 - 4.05) | 3.86 |
| *NR3C2* |  |  |  |  |  |  |  |
| Disruptive | 3 | 3 | 0.000022 | 0 | 0.000000 |  | -0.26 |
| Splice site variant | 1 | 1 | 0.000007 | 0 | 0.000000 |  | 0.00 |
| *KCNJ5* |  |  |  |  |  |  |  |
| Disruptive | 10 | 16 | 0.000119 | 12 | 0.000181 | 1.51 (0.70 - 3.27) | 0.50 |
| Splice site variant | 1 | 1 | 0.000007 | 0 | 0.000000 |  | 0.00 |
| *CACNA1H* |  |  |  |  |  |  |  |
| Disruptive | 56 | 84 | 0.000625 | 35 | 0.000529 | 0.86 (0.58 - 1.28) | -0.34 |
| Splice site variant | 21 | 27 | 0.000201 | 14 | 0.000212 | 1.10 (0.57 - 2.13) | 0.06 |
| *CACNA1D* |  |  |  |  |  |  |  |
| InDel | 9 | 1360 | 0.010111 | 860 | 0.013006 | 1.30 (1.19 - 1.42) | 8.10 |
| Disruptive | 12 | 8 | 0.000059 | 4 | 0.000060 | 1.11 (0.32 - 3.81) | 0.00 |
| Splice site variant | 6 | 5 | 0.000037 | 4 | 0.000060 | 1.80 (0.47 - 6.91) | 0.31 |
| *CLCN2* |  |  |  |  |  |  |  |
| Synonymous | 180 | 1167 | 0.008676 | 523 | 0.007910 | 0.89 (0.80 - 0.99) | -1.45 |
| 3 prime UTR | 34 | 319 | 0.002372 | 125 | 0.001890 | 0.78 (0.63 - 0.96) | -1.78 |
| Disruptive | 41 | 87 | 0.000647 | 36 | 0.000544 | 0.85 (0.57 - 1.26) | -0.40 |
| Splice site variant | 14 | 14 | 0.000104 | 8 | 0.000121 | 1.05 (0.43 - 2.57) | 0.09 |
| *ATP1A1* |  |  |  |  |  |  |  |
| Synonymous | 201 | 5833 | 0.043367 | 3493 | 0.052826 | 0.95 (0.91 - 1.00) | -1.48 |
| Disruptive | 7 | 4 | 0.000030 | 4 | 0.000060 | 1.95 (0.47 - 8.10) | 0.34 |
| Splice site variant | 1 | 1 | 0.000007 | 0 | 0.000000 |  | 0.00 |
| *ATP2B3* |  |  |  |  |  |  |  |
| Disruptive | 9 | 226 | 0.001680 | 74 | 0.001119 | 0.91 (0.78 - 1.06) | -0.64 |
| Splice site variant | 2 | 2 | 0.000015 | 0 | 0.000000 |  | 0.00 |
| *KDM1A* |  |  |  |  |  |  |  |
| 3 prime UTR | 28 | 43 | 0.000320 | 34 | 0.000514 | 1.61 (1.01 - 2.56) | 1.41 |
| Disruptive | 32 | 21 | 0.000156 | 20 | 0.000302 | 1.93 (1.05 - 3.58) | 1.35 |
| Splice site variant | 7 | 8 | 0.000059 | 3 | 0.000045 | 0.79 (0.20 - 3.08) | 0.00 |

## Discussion

These analyses provide a broad overview of some impacts of very rare genetic variants in a large sample broadly representative of the population. It should be pointed out that the analytic approach used assumes that all variants in gene have the same direction of effect. Since any random variant is more likely to impair the function of a gene than to enhance it, weighted burden analysis is not expected to detect effects of gain of function variants because these are likely to be swamped by other variants. Likewise the method is expected to be relatively insensitive at detecting variants which have a purely recessive effect. Of course it is quite possible that the effect of complete loss of function of one gene may be modified by functional variants affecting the other copy of the gene but such complex heterozygous effects are difficult to detect. Additionally, a population sample differs from a specially recruited case-control sample in that it is not enriched for the phenotype in question and hence will have less power to detect either rare recessively acting variants or extremely rare variants with a dominant effect. Thus, we expect that there may well be important rare variant effects in addition to the ones which this study has highlighted.

Another contrast with specifically defined case-control studies is that the phenotype needs to be derived from measures which are provided and in order to improve power it is desirable to use information which is available for a large number of participants. The phenotype used here attempts to reflect a clinical diagnosis of hypertension but of course will not do this as accurately as could be achieved in a specifically ascertained sample. Some participants may be on antihypertensive medication for indications other than hypertension, some may have undiagnosed hypertension and for some the diagnosis may simply be incorrect. Measured blood pressure itself was not used to inform the phenotype, in part because it might reflect blood pressure on medication and in part because single measurements may be less informative than indirectly relying on the clinical decisions which have fed into assigning a diagnosis or starting a prescription. The advantages of having a large sample to some extent balance the disadvantage of a less accurate phenotype.

When considering the contribution of risk within the population, we should note that the variants involved are rare. Although a MAF threshold of 0.01 was used, the majority of variants analysed are very much rarer than this and for the variant categories with the most severe consequences the cumulative frequency of variants in the category is also low. This means that few subjects will carry more than one variant with a severe consequence and we can say that the mean count of variants of a particular category is a good approximation for the proportion of the subjects who carry a variant of that category.

The result for *DNMT3A* seems unlikely to be due to chance since it would remain significant at p = 0.00016 even after correction for the number of genes tested. The results show that LOF variants in this gene are associated with nearly doubling the OR for hypertension and are present in about 1 in a thousand people, while a slightly larger number will have a variant annotated as probably damaging by PolyPhen which moderately increases hypertension risk. *DNMT3A* is a DNA methyltransferase and nonsynonymous variants in it have been reported as causes of two different syndromes, Tatton-Brown-Rahman syndrome with overgrowth and intellectual disability and Heyn-Sproul-Jackson syndrome with microcephalic dwarfism, neither of which has hypertension as part of the phenotype (Heyn et al., 2019; Tatton-Brown et al., 2014). Missense variants in *DNMT3A* have also been reported in autism spectrum disorder and mice with Dnmt3a haploinsufficiency produced by heterozygous deletion of exon 19 have increased body weight and some behavioural alterations (Christian et al., 2020). In a series of 210 patients with an overgrowth syndrome similar to Sotos syndrome but with no *NSD1* mutation, four had de novo nonsynonymous mutations in *DNMT3A* and two had stop variants (Tlemsani et al., 2016). One of the stop variants was inherited from a normal mother in whom it was thought a somatic mutation had occurred and for the other the father’s DNA was not available. Given the frequency of LOF variants in *DNMT3A* observed in our samples, it seems possible that the observation of two patients with stop variants was coincidental. Thus it may be that, while certain specific nonsynonymous variants can cause severe phenotypes, generally reduced functioning of *DNMT3A* does not cause marked problems but is associated with increased risk of hypertension.

Although *DNMT3A* variants have not previously been reported to be associated with hypertension, as reviewed recently these findings are consistent with a body of results relating to the histone lysine demethylase LSD1, which together implicate LSD1 hypofunction in salt-sensitive hypertension (Hirohama and Fujita, 2019). In our data disruptive variants in *KDM1A*, the gene for LSD1, are associated with an OR of 1.9 but they are too rare for firm conclusions to be drawn. DNMT3A methylates DNA conditional on the associated H3K4 residue being unmethylated and LSD1 accomplishes this demethylation, meaning that reduced function of LSD1 is expected to reduce DNMT3A-dependent DNA methylation (Hashimoto et al., 2010; Shi et al., 2004). In mice, *Dnmt3a* deficiency, which can be produced by knockdown or as a consequence of foetal exposure to dexamethasone or low protein maternal diet, has been shown to lead to reduced methylation of the gene for angiotensin receptor type 1a, *Agtr1a*, leading to increased *Agtr1a* expression and salt-induced hypertension (Kawakami-Mori et al., 2018). Thus, a consistent picture emerges that reduced DNMT3A activity can increase risk for hypertension, whether this is due to genetic variants in *DNMT3A* itself, LSD1 hypofunction or the uterine environment.

Although *FES* (SLP = 6.10) only just reaches criteria for exome-wide significance, confidence in this result is somewhat increased by the fact that a nearby SNP, rs2521501, shows robust evidence for association (Ehret et al., 2011). However the mechanisms by which it might influence hypertension risk are unclear as it codes for a tyrosine kinase which is involved in various signalling pathways and which may have a role in haematopoiesis and regulating the innate immune response (Zirngibl et al., 2002).

The results for *GUCY1A1* (SLP = 5.54) and *GUCY1B1* (SLP = 3.92) are more compelling, given that they code for two different subunits of the same protein, soluble guanylate cyclase, and given that recessively acting variants in *GUCY1A1* have previously been reported in cases of moyamoya disease with hypertension (Wallace et al., 2016). Soluble guanylate cyclase is responsible for detecting NO signalling in order to produce vasodilation and other responses, and the central role of this pathway in the control of blood pressure is well-established from animal studies while guanylate cyclase stimulators have been developed as treatments for pulmonary hypertension (Buys and Sips, 2014). The findings reported here are the first to directly demonstrate that impaired functioning of either of these genes represents a risk factor for systemic hypertension in the general population, with nearly 1 in a thousand people carrying a LOF variant in one of them which approximately doubles the OR for hypertension.

The results for *NPR1* (SLP = 5.14) represent a replication of the previously reported findings and confirm that, although certain nonsynonymous variants such as rs61757359 may be associated with reduced blood pressure, in general variants which impair the functioning of this gene increase the risk of hypertension (Liu et al., 2016; Vandenwijngaert et al., 2019). Around 1 in 500 people carries a variant annotated by PolyPhen as probably damaging and overall such variants are associated with a modest increase in risk with OR = 1.32 (1.05 - 1.66).

*SMAD6* (SLP = 4.10) has a role in signalling pathways and has not previously been clearly implicated in hypertension risk although a recent report describes how exome sequencing of 37 children with renovascular hypertension revealed a frameshift variant classified as likely pathogenic variant in *SMAD6* in one patient (Viering et al., 2020). *SMAD6* variants are known to predispose to cardiovascular malformations including bicuspid aortic valve related aortopathy (Gillis et al., 2017; Luyckx et al., 2019; Tan et al., 2012). The results reported here suggest that LOF variants in this gene may have a moderate effect on increased risk of hypertension in the general population.

Recessively acting variants in *IFT172* (SLP = 3.39) can cause ciliopathies and there is a report of a child with compound heterozygous variant who presented with growth retardation and subsequently developed retinopathy, metaphyseal dysplasia and, at the age of 11, hypertension (Lucas-Herald et al., 2015). However there does not seem to be other evidence to implicate *IFT172* in hypertension risk so this result would require replication in other samples.

*CSK* (SLP = 3.37) is located in the 15q24 locus which, as reviewed recently, is implicated by multiple GWAS for hypertension (Lee et al., 2016). Following up the results of eQTL analyses, these authors demonstrated that mice with gene-silencing or haploinsufficiency of *Csk* had increased blood pressure and showed that this effect could be moderated by PP2, an inhibitor of Src. Although the results reported here are not formally significant after correction for multiple testing, the additional support provided by these GWAS findings and functional studies does suggest that variants in *CSK*, in particular those annotated as deleterious by SIFT, might be a risk factor for hypertension.

The results for *DBH* (SLP = −3.40) provide further support for the previously reported findings that variants in this gene are associated with blood pressure (Liu et al., 2016). In particular, the results suggest that variants annotated as deleterious by SIFT are on average associated with a slightly reduced risk of developing hypertension. Although these variants are individually rare, about 1 person in 20 carries one of them.

*AGTR1* (SLP = −3.77) codes for a receptor for angiotensin II so it seems very plausible from a biological point of view that variants impairing its function might be protective against hypertension in spite of the fact that no association with common variants has been detected (Ji et al., 2017). The results suggest that very rare gene disruptive variants can about halve the OR for hypertension.

*ZYX* (SLP = −3.83) is potentially of interest because it codes for zyxin, the protein responsible for sensing stretch in endothelial cells and vascular smooth muscle cells, as occurs in hypertension, and mediating their response to this by changing the expression of other genes (Ghosh et al., 2015). The findings reported here suggest that impaired functioning of this gene may reduce risk of hypertension.

*PREP* (SLP = −5.03) narrowly fails to meet conventional criteria for exome-wide significance but is clearly of interest because its product, prolyl endopeptidase, also known as prolyl oligopeptidase or post-proline cleaving enzyme, has recently been shown to be responsible for converting circulating angiotensin II to angiotensin-(1-7) in the circulation and in lungs, a process which is largely independent of ACE2, which carries out this conversion in the kidney (Serfozo et al., 2020). The results we report suggest that rare, functional variants in *PREP* are protective against hypertension but it is not clear which categories of variant are responsible and although LOF variants are commoner in controls they are too rare for conclusions to be drawn. In mice, loss of this gene results in reduced ability to metabolise ACE2 and hence a more prolonged systemic hypertensive response to exogenously administered ACE2 (Serfozo et al., 2020). While it is not obvious why impaired functioning of this gene might be protective against hypertension these findings do seem worthy of further exploration.

The finding that an inframe deletion in *CACNA1D*, rs72556363, is associated with increased of hypertension is consistent with reports that very rare germline and somatic variants in this gene can result in aldosterone-producing adenomas and primary aldosteronism (Scholl et al., 2013). Although the variant varies in frequency between populations, essentially being restricted to those with European ancestry, this result does not seem likely to be due to an artefact of population stratification because the frequency in controls is similar to that reported in non-Finnish Europeans whereas the frequency in cases is even higher. It seems to represent a modest risk factor for hypertension without producing severe hyperaldosteronism, which is found in about 1% of subjects with European ancestry.

Overall, these analyses provide an overview of some of the impacts rare, coding variants may have on the risk of hypertension in the general population. The validity of some novel findings will become clearer when exome sequence data is released for the remaining 300,000 UK Biobank participants or if they can be tested in other samples or followed up in functional studies. All the variants implicated are very rare and in view of their effect sizes arguably do not make an important contribution to risk from a public health point of view. Nor are they probably helpful as individual measures of risk, partly because they can only be detected by sequencing. Although it may be reasonable to assume that LOF variants within a given gene will tend to have a similar effect on phenotype, the same cannot be said of nonsynonymous variants and even within a given category the effect of such variants is likely to be vary considerably. Thus, for variants which are individually extremely rare it is in general not possible to make a clear interpretation regarding their likely effect. The main value of these findings is probably in highlighting genes and biological pathways of relevance in order ultimately to inform improved therapeutic approaches.

## Data Availability

The raw data is available on application to UK Biobank. Detailed results with variant counts cannot be made available because they might be used for subject identification. Scripts and relevant derived variables will be deposited in UK Biobank. Software and scripts used to carry out the analyses are also available at https://github.com/davenomiddlenamecurtis.

## Conflicts of interest

The author declares he has no conflict of interest.

## Preprint availability

A preprint describing this work has been posted at medRxiv with doi 10.1101/2021.02.10.21251503 (Curtis, 2021d).

## Acknowledgments

This research has been conducted using the UK Biobank Resource. The author wishes to acknowledge the staff supporting the High Performance Computing Cluster, Computer Science Department, University College London. This work was carried out in part using resources provided by BBSRC equipment grant BB/R01356X/1. The author wishes to thank the participants who volunteered for the UK Biobank project.

## Ethics statement

UK Biobank had obtained ethics approval from the North West Multi-centre Research Ethics Committee which covers the UK (approval number: 11/NW/0382) and had obtained informed consent from all participants. The UK Biobank approved an application for use of the data (ID 51119) and ethics approval for the analyses was obtained from the UCL Research Ethics Committee (11527/001).

## Author contributions

DC carried out the analyses and prepared the manuscript.

## Notes

### Competing Interest Statement

The authors have declared no competing interest.

### Funding Statement

No external funding was received.

### Summary of Updates

There was a typo in the name of CACNA1D

